# Work-related musculoskeletal disorders and associated factors among cleaners of health institutions in Gondar town, Northwest Ethiopia: an institution based cross-sectional study

**DOI:** 10.1101/2021.12.06.21266337

**Authors:** Jemal Suleyman, Asmare Yitayeh Gelaw

**Affiliations:** Department of physiotherapy, School of Medicine, College of Medicine and Health Sciences, University of Gondar, Gondar, Ethiopia

**Keywords:** cleaners, health institution, musculoskeletal disorders

## Abstract

**Background:** Musculoskeletal disorders are growing public health problems both in developed and developing countries including Ethiopia. However, its prevention and control has not yet received due attention. This study aimed to determine.

the prevalence and associated factors of musculoskeletal disorders musculoskeletal disorders among cleaners of health institutions in Gondar town, Northwest Ethiopia

**Method:** An institutional based cross-sectional study was conducted, from April to May 2016 in all health institutions of Gondar town. All the available cleaners of health institutions were taken as study participants. Data were collected by face-to-face interview technique after verbal informed consent. Additionally, weight and height of participants were measured following standard procedures. Data were collected by trained physiotherapists and then entered into a computer using Epi Info version 3.5.3 and exported to SPSS version 20 for analysis. Descriptive statistics was performed to describe the data in percentage and mean. Multiple logistic regressions were fitted and Odds ratios with 95% confidence intervals were calculated to identify associated factors.

**Results:** A total of 242 participants were included in this study. The majority of the study participants were females (79.3%) and between 25-44 age group (65.3%). Two hundred 0ne (83.1%) of the respondents reported that they had pain in at least one of the body parts in the previous 12 months. Of the nine body parts examined, neck (76%), upper back (40.5%) and lower back (45%) were the most frequently body parts reported to exhibit pain. Job status (AOR = 2.71, 95%CI; 1.37-5.36), and static work habit (AOR = 2.71, 95%CI; 1.37-5.36), were factors associated with musculoskeletal disorders.

**Conclusion:** There is a high prevalence of musculoskeletal disorders among cleaners in the health institutions of Gondar Town. Job status and static work habit were the significant associated factors. Hence, we recommend the design and implementation of institution based screening programs for musculoskeletal disorders

## Statement of the Problem

Work-related musculoskeletal disorders are a major health issue in many occupations all over the world(1,2).Extensive research has been conducted on these musculoskeletal problems in different occupational groups such as health care professionals, building construction workers, office workers, bus drivers and sewing machine operators(3–6)

The cleaning services in health institutions represent one of the most dynamic occupations which expose cleaners to diverse occupational hazards including musculoskeletal disorders. A broad range of cleaning activities, from sweeping and vacuuming to disposing of waste or cleaning toilets is performed in health institutions. These Cleaning activities involve both dynamic and static muscular work with the use of various pieces of manual equipment. Cleaning work is generally labor intensive and involves repetitive movements, awkward positioning, and stretching which necessitate high cardio respiratory and musculoskeletal demands(7).

Factors contributing for the development of WMDs among cleaners in health institutions are wide spread and mostly occupational in origin(1,7–11). Sociodemographic factors (age and sex) and psychosocial factors are also important underlying factors for the progress of WMDS among cleaners in health institutions(10,12,13)

Ever since few studies have investigated work related musculoskeletal disorders among cleaners of health institutions both in developed and developing countries, there is limited information available for the prevalence and contributing factors of WMDs among cleaners of sub-Sahara African countries.. A knowledge gap therefore exists in scientific literature on the prevalence of WMDs and associated factors among cleaners of health institutions.).Therefore, this study aimed to assess the prevalence of WRMSDs and associated factors among cleaners of health institutions working at Gondar town.

### 2.2. Method

### 2.1. Study design and setting

An institutional based cross-sectional study was undertaken in health institutions of Gondar town to assess the prevalence and associated factors of WRMSDs among cleaners of health institutions in Gondar town from April to May, 2016. It was conducted in health institutions of Gondar town. Gondar town is located 750kilometers from Addis Ababa, the Ethiopian capital. The town has 12 “kifleketemas” (sub-towns) and 21 kebeles(the smallest administrative units in Ethiopia) and is among the ancient and densely populated cities in Ethiopia having 206,987people according to 2007 Ethiopian Central Statistical Agency (CSA) office report (14,15).

It has one referral hospital (UOG hospital), eight local governmental health centers and 42 private health institutions.

### 3.3. Source and study population

All cleaners working in health institutions of Gondar town were considered as source population. All cleaners who were available in all HIs of Gondar town during the data collection period were considered as study population. The total number of cleaners in the health institutions was 242.The whole population was taken for the study due to being small population size.

### 3.7. Inclusion and Exclusion criteria

#### 3.7.1. Inclusion criteria

All cleaners who were available in all HIs of Gondar town during data the collection

#### 3.7.2. Exclusion criteria

All cleaners under 1 year work duration

### 3.8. Operational definitions

**Work related musculoskeletal disorders**: perceived pain, ache or discomfort in any part of body segments caused, aggravated or exacerbated by work place exposures.

**Body segments**: neck, shoulder, upper back, lower back, hip /thigh, knee/leg and ankle/foot, wrist /hand

### 3.9. Data collection tools and procedure

A standardized structured questionnaire adapted from the Nordic musculoskeletal questionnaire, with modification to suit the local context consisting of both closed and open ended questions was used to collect the data. Data were collected using a combination of a structured questionnaire and measurements of weight and height. Data collectors were six physiotherapists supervised by investigators. Training and practical demonstrations on interview techniques and measurement procedures were given to data collectors for two consecutive days. The questionnaire was pretested on 5% of the study participants found outside of the study area and modifications were made on the basis of the findings. After completing the interview, the participant’s height and weight were measured and recorded by interviewers. Weight measuring scales were checked and adjusted atzero level between each measurement and height was measured following the standard steps.

### 3.10. Data management and analysis

Data were coded and entered into Epi Info version 7 and exported to SPSS version 16 for analysis. Overall proportion with 95%confidence interval (CI) had been calculated to determine the prevalence. For testing of significance, categorical data were compared using chi-square test. Associated factors of WMDs were determined by using descriptive analysis. Multicollinearity test was also checked to assess the correlation between age and professional experience, height and BMI, sex and pregnancy. Logistic regression model had been used to assess the association of independent variables with WMDs. OR with 95% (CI) for risk indicators was calculated. The statistical tests were considered significant at a level <5% (<0.05).

## Ethical considerations

The study was done after ethical clearance was secured from the Ethical Review Committee of College of Medicine and Health sciences, University of Gondar. Informed consent was obtained from study participants after being informed in detail about the objective, purpose, benefits and risks of the study. Appropriate measures were taken to assure confidentiality of information both during and after data collection. Identified subjects with WMDs during data collection time were given advice or referred, for further care, to physiotherapy

## 5. Result

A total of 242 cleaners (95.7% response rate) from health institutions of Gondar town were included in this study. The majority of participants (79.3%) were females. The mean age was 40.5± 8.49 years.

. The mean and standard deviation age of participants was 40.55 respectively. The majority of the participants were in the 25-44 age group, 158 (65.3%).All the participants were from three types of health institutions, namely UOG referral Hospital (69.8%), LGHIs (10.7%), private HIs (19.4%).The majority of the participants had working experience of less than two years. Table 1 shows the detail of the socio-demographic characteristics.

**Table 1:**
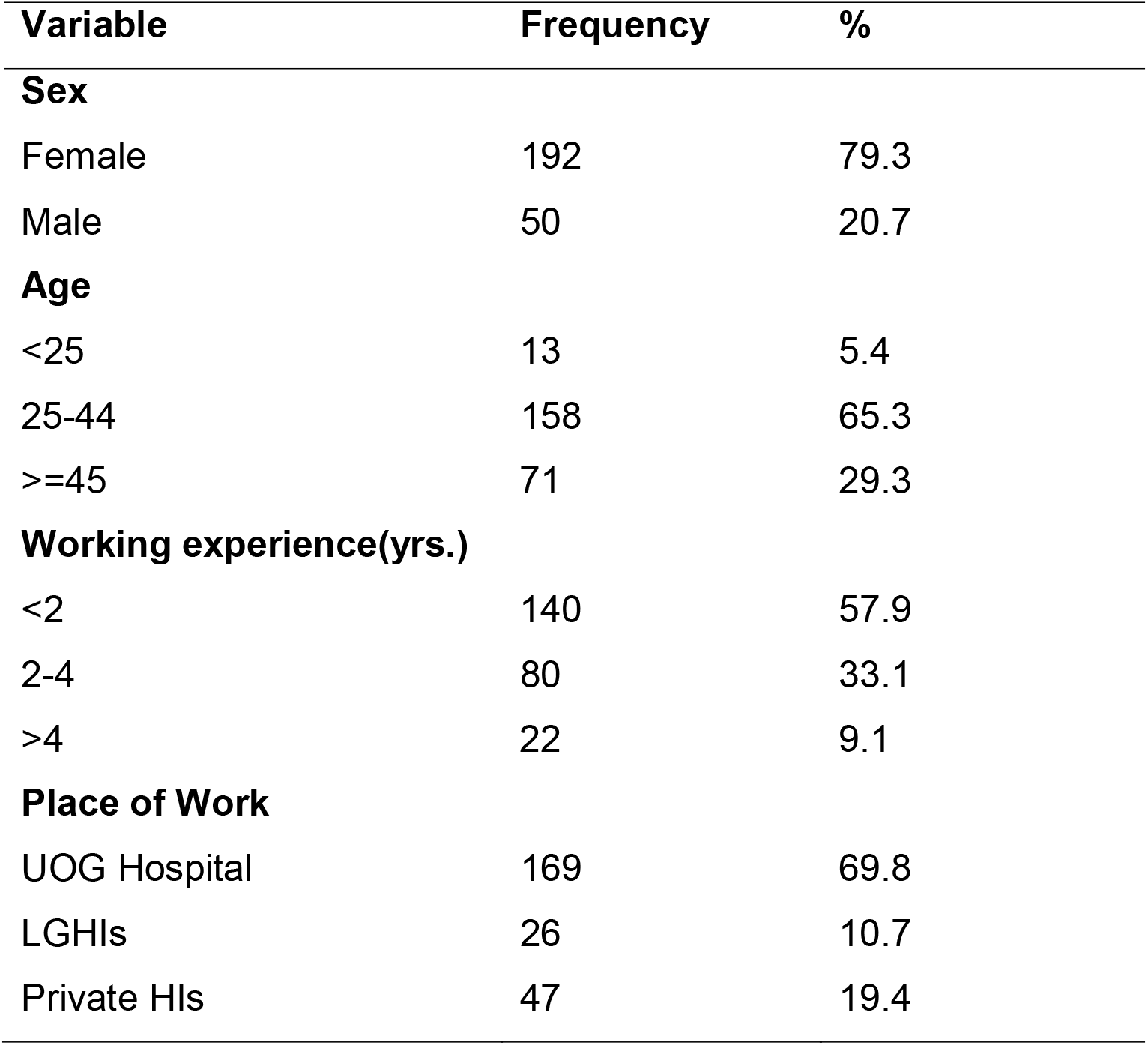
Distribution of cleaners by socio-demographic characteristics in Gondar town, 2016.(n=242)

### 5.2. Work -related and Psychosocial Characteristics of the Respondents

The majority of the participants were in the group of 8 hours duration of work per day 156 (64.5%). Participants with permanent job status were 223 (92.1%). Majority of the respondents reported that they had no comfortable work schedule194 (80.2%). Majority of the respondents reported that they had no time break 160 (66.1%). Regarding job satisfaction 220 (90.9%) reported that they were not satisfied with their job. Table 2 shows the detail of these factors.

**Table2.** Work related and psychosocial characteristics of cleaners in Gondar town, 2016 (n=242)

| **Variable** | **Frequency** | **%** |
| --- | --- | --- |
| **Job status** |  |  |
| Permanent | 223 | 92.1 |
| Temporary | 19 | 7.9 |
| **Comfortable work schedule** |  |  |
| Yes | 48 | 19.8 |
| No | 194 | 80.2 |
| **Time break** |  |  |
| Yes | 82 | 33.9 |
| No | 160 | 66.1 |
| **Job satisfaction** |  |  |
| Yes | 22 | 9.1 |
| No | 220 | 90.9 |
| **Job control** |  |  |
| Yes | 19 | 7.9 |
| No | 229 | 92.1 |
| **Duration of work/day (hrs.)** |  |  |
| <8 | 50 | 20.7 |
| 8 | 156 | 64.5 |
| >8 | 36 | 14.9 |

**Table3.** Results of binary and logistic regression for the associated factors of WRMSDs among cleaners in Gondar town, Northwest Ethiopia, 2016 (n=242)

| Variables | Musculoskeletal Disorders |  | COR (95% CI) | AOR (95% CI) |
| --- | --- | --- | --- | --- |
|  | Yes | No |  |  |
| **Job status** |  |  |  |  |
| Permanent | 201 | 41 | 4.41(2.026,14.245)* | 11.17(3.293,37.85)** |
| Temporary | 10 | 9 | 1 | 1 |
| **Static work habit** |  |  |  |  |
| Yes | 33 | 16 | 0.31(0.148,0.637)* | 0.27(0.107, 0.689)** |
| No | 168 | 25 | 1 | 1 |
| **Comfortable work schedule** |  |  |  |  |
| Yes | 173 | 21 | 5.84(2.833,12.223)* | 2.21(0.799, 6.097) |
| No | 28 | 20 | 1 | 1 |

\*: statistically significant at $p < 0.05$ , \*\*: statistically significant at $p \leq 0.01$ , OR: odd ratio, CI: confidence interval, 1: reference

### 5.3. Prevalence of musculoskeletal pain among cleaners

Majority of the participants 201 (83.1%) reported that they had pain at least in one of the body segments in the previous 12 months. The most prevalent body segments reported to exhibit pain among the participants were neck (76%), lower back (45%) and upper back (40.5%), respectively. The following figure shows the prevalence of pain in the nine body parts.

**Figure 2:**
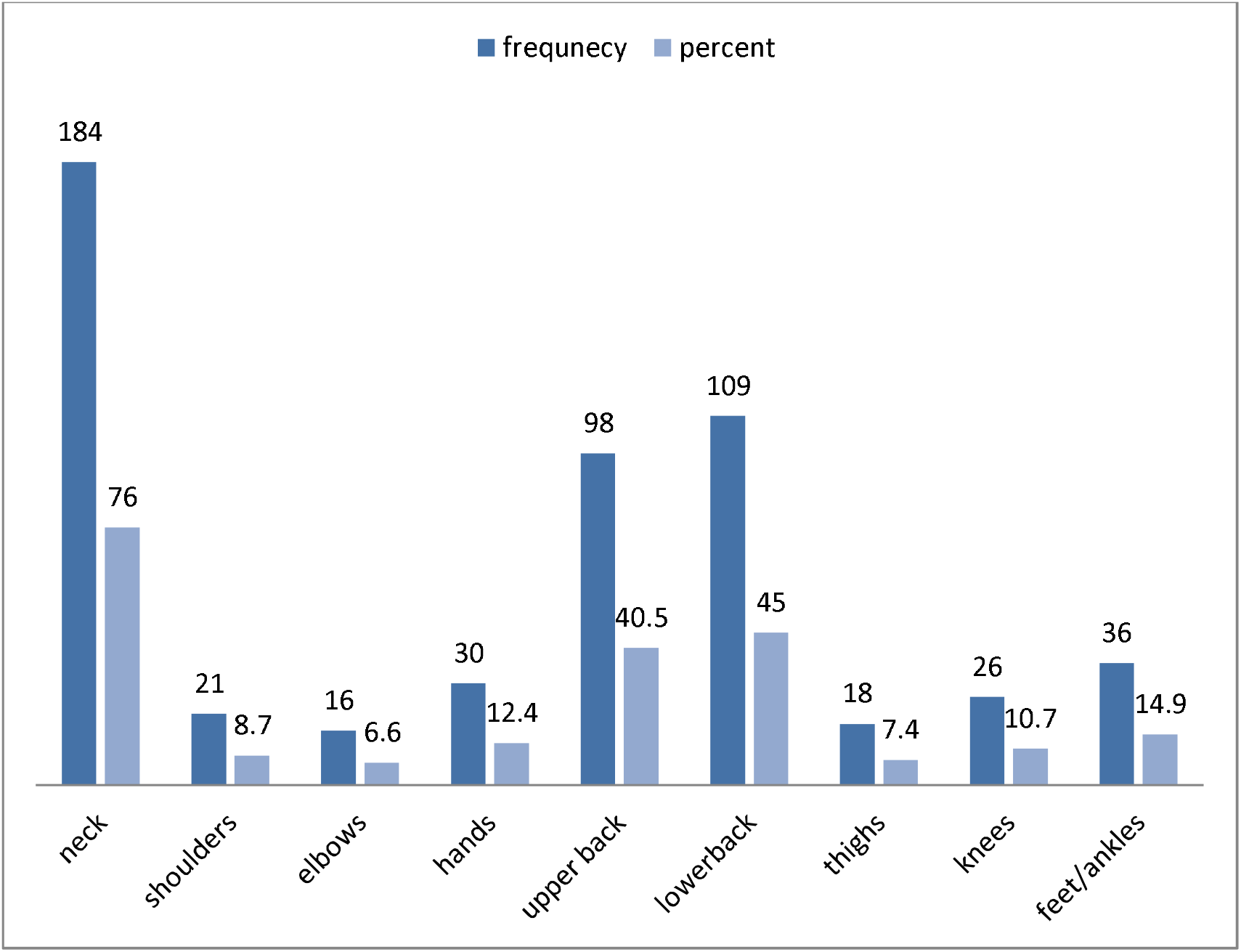
Prevalence of WRMSD on each body parts.

### 5.4. Factors associated with WRMSDs among the respondents

Bivariate and multivariate logistic regression analyses were used to identify factors associated with WRMSDs among cleaners of HIs in Gondar town. The crude (COR) and adjusted odds ratios (AOR) together with their corresponding 95% confidence interval were computed. Job status, static work habit, and comfortable work schedule were significantly associated variables with WRMSDs.

After fitting these associated variables in to multiple logistic regressions, job status and static work habit were independently associated with WRMSDs.

Job status was found to be significantly associated WRMSDs. Those who had full time were 11.7 (AOR= 11.17, 95 % CI(3.29, 37.85)) times more likely develop WRMSDs as compared to those of part time job status.

Static work habit was also found to be significantly associated to WRMSDs. The result showed those who had no static work habit were 70% times less likely to develop MSD than those with static work habit (AOR=0.271, 95%CI;(0.107, 0.689)).

## 6. Discussion

The purpose of this study were to determine the prevalence of musculoskeletal disorders musculoskeletal disorders and to identify the factors associated with musculoskeletal disorders among cleaners in health institutions of Gondar town in the previous 12 months. In this study, female workers made up the majority (79.3 %) of the cleaners. This was similar with the study done in Norway (82.8%)(16)and Nigeria (92.4%)(16).

This study found that about 83.1% of the participating cleaners complained of musculoskeletal disorders. This was close to the prevalence reported in Taiwan 90%(16). However, it was higher if compared with the reported prevalence of musculoskeletal disorders musculoskeletal disorders among cleaners in Norway56%(16). The higher 12-months prevalence found in the current study suggests that cleaners’ practice in Ethiopia highly predisposes to musculoskeletal disorders. This may be a reflection of the work settings under which cleaners practice in Ethiopia.

The neck was the most affected body part with a percentage of 76% in this study. This was higher prevalence rate reported in comparison with the previous study as in Germany (49%)(16).The difference may be due to work setting difference between developing and developed country. The causes of neck disorders may be due to performing repetitive movement lifting too much weight or lifting improperly.

The lower back was the second most affected body part with a percentage of 45% in this study. This was slightly higher prevalence rate reported in comparison with the previous study conducted in Taiwan (37.8%)(16).The differences may be due to difference of work setting in developing and developed country in which the developed countries may use technologies that reduce stress for the back.

The upper back was the third most affected body part with a percentage of 40% in this study. This was higher prevalence rate reported in comparison with the previous study conducted in Norway (21%)(16). The differences may be due to difference of work setting in developing and developed country in which the developed countries may use technologies that reduces stress to the back.

Job status was significantly associated with WRMSDs. The permanent workers had higher prevalence of WRMSDs than the temporary workers (AOR= 11.17, 95 % CI (3.29, 37.85)). This may be due to the work load happened due to daily work. This finding was supported by a study conducted in Iran(16), in Taiwan(16) and China(16).

Static work habit was also significantly associated with WRMSDs (AOR=0.271, 95%CI;(0.107, 0.689)).This may due to the fact that more stress is applied due to working in a fixed position so that they feel pain. This finding was supported by a study conducted in America(16).

## 7. Limitations of the study

The strength of this study is somewhat weakened because both exposure and musculoskeletal discomfort were ascertained by self-report, instead of objective documentation or medical records. Some of the included variables were not measured according to a standard measuring category.

## 8. Conclusion

This study interviewed 242 cleaners in health institutions of Gondar town to evaluate the prevalence and associated factors of WRMSDs. The prevalence of WRMSDs among cleaners of HIs in Gondar town was high. Among the nine body parts investigated, neck, upper back and low back were the most common body parts with musculoskeletal complaints. Job status and static work habit were identified as factors that affect WRMSDs.

## 9. Recommendation

Based on the findings of the study, the following points are recommended to prevent and manage the magnitude of WRMSDs among cleaners of HIs in Gondar town.

### For researchers

✓ Further study with standardized measurement of important variables and data collection aided with medical records and other materials
✓ Further study with wide geographical population
✓ Further study with different study design such as RCT.

**For clinicians**-they are expected to giveadvice for cleaners related to work posture.

**For employers**-they are expected to consider timing of work and the way cleaners work in the organization

**For policy makers-**enforcing the establishment of preventive and curative aspects for WRMSDs

## Data Availability

all data produced in the present work are contained in the manuscript

## Notes

### Competing Interest Statement

The authors have declared no competing interest.

### Funding Statement

this study was funded by university of Gondar

### Author Declarations

Ethical clearance was obtained from the Ethical Review Committee of school of medicine, UOG.

